# NON-ENDOSCOPIC ESOPHAGEAL SAMPLING DEVICE AND BIOMARKER PANEL FOR DETECTION OF BARRETT’S ESOPHAGUS (BE) AND ESOPHAGEAL ADENOCARCINOMA (EAC)

**DOI:** 10.1101/2023.06.06.23291048

**Authors:** Helen R. Moinova, Suman Verma, John Dumot, Ashley Faulx, Prasad G. Iyer, Marcia Irene Canto, Jean S. Wang, Nicholas J. Shaheen, Prashanthi N. Thota, Lishan Aklog, Joseph E. Willis, Sanford D. Markowitz, Amitabh Chak

## Abstract

**BACKGROUND:** We previously reported an encapsulated balloon (EsoCheck^TM^, EC), which selectively samples the distal esophagus, that coupled with a two methylated DNA biomarker panel (EsoGuard^TM^, EG), detected Barrett’s esophagus (BE) and esophageal adenocarcinoma (EAC), with a sensitivity and specificity of 90.3% and 91.7%, respectively. This previous study utilized frozen EC samples.

**AIM:** To assess a next generation EC sampling device and EG assay that utilizes a room temperature sample preservative to enable office-based testing.

**METHODS:** Cases with nondysplastic (ND) and dysplastic (indefinite=IND, low grade dysplasia = LGD, high grade dysplasia = HGD) BE, EAC, junctional adenocarcinoma (JAC) and controls with no intestinal metaplasia (IM) were included. Nurses or physician assistants at six institutions, who were trained in EC administration, delivered the encapsulated balloon per orally and inflated it in the stomach. The inflated balloon was pulled back to sample 5 cm of the distal esophagus, then deflated and retracted into the EC capsule to prevent sample contamination from proximal esophagus. Nextgen EG sequencing assays performed on bisulfite-treated DNA extracted from EC samples determined levels of methylated Vimentin (mVIM) and methylated Cyclin A1 (mCCNA1) in a CLIA-certified laboratory, blinded to patients’ phenotypes.

**RESULTS:** A total of 243 evaluable patients – 88 cases (median age 68 years, 78% men, 92% white) and 155 controls (median age 57 years, 41% men, 88% white) – underwent adequate EC sampling. Mean time for EC sampling was just over 3 minutes. The cases included 31 NDBE, 16 IND/LGD, 23 HGD, and 18 EAC/JAC. Thirty-seven (53%) of the non-dysplastic and dysplastic BE cases were short-segment BE (SSBE; < 3 cm). Overall sensitivity for detecting all cases was 85% (95% CI= 0.78-0.93) and specificity was 85% (95% CI=0.79-0.90). Sensitivity for NDBE was 84% (n=37). The EC/EG test detected 100% of cancers.

**CONCLUSION:** The next-generation EC/EG technology has been both successfully updated to incorporate a room temperature sample collection preservative and successfully implemented in a CLIA certified laboratory. When performed by trained personnel, EC/EG detects non-dysplastic BE, dysplastic BE, and cancer with high sensitivity and specificity, replicating the operating characteristics of the initial pilot study of this technology. Future applications utilizing EC/EG to screen broader populations at risk for developing cancer are proposed.

**SIGNIFICANCE:** This multi-center study demonstrates the successful performance of a commercially available clinically implementable non-endoscopic screening test for BE in the U.S., as recommended in the most recent ACG Guideline and AGA Clinical Update. It transitions and validates a prior academic laboratory-based study of frozen research samples over to a CLIA laboratory, one that also integrates a clinically practical room temperature method for sample acquisition and storage, enabling office-based screening.

## Introduction

Esophageal cancer is the 8^th^ most-commonly diagnosed cancer globally, ranking number six in global cancer-related mortality[1, 2]. Esophageal adenocarcinoma (EAC) is the predominant subtype of esophageal cancer in the US, Northern and Western Europe, and Australia, and the incidence of EAC has more than quadrupled in the past thirty years [3–6]. The prognosis for EAC patients remains poor, with a less than 20% survival at 5 years [6, 7]. Early diagnosis is the most important factor in determining the survival of EAC patients [2], but most EACs are diagnosed at a later stage, when treatment options are limited, therefore early detection would have a major impact on the survival.

Barrett’s esophagus (BE), a pre-malignant intestinal-type columnar metaplasia that replaces the normal squamous mucosa of the distal esophagus, is the only known precursor for EAC, but its detection currently requires performing esophagogastroduodenoscopy (EGD). Due to the high cost of EGD and the lack of a randomized controlled trial demonstrating cost effective reduction in EAC, endoscopy screening for BE has not been routinely recommended [8, 9]. Thus, in approximately 95% of cases of EAC, the presence of the antecedent BE remains undetected and unknown [10].

We previously reported that an encapsulated balloon device (EsoCheck ^TM^, EC), which selectively samples the distal esophagus, coupled with a two methylated DNA biomarker panel (EsoGuard^TM^, EG), could detect BE and EAC, with a sensitivity and specificity of 90.3% and 91.7%, respectively[11]. The previously-published version of the assay required EC samples to be immediately frozen and kept at -70C until DNA extraction. In order to make the assay practically-useable for widespread office-based testing, we have developed a buffer that preserves the DNA methylation signature in samples collected and stored at ambient temperature.

The EsoGuard technology has been licensed to Lucid Diagnostics, and the assay transferred from an academic research laboratory over to Lucid’s CLIA-compliant clinical lab. The aim of this study was to perform a case control study to: i) evaluate the performance of the EsoGuard assay in this updated format as run at the Lucid clinical laboratory on samples collected in a room temperature in preservative buffer, and ii) confirm the sensitivity and specificity of the assay in a larger independent patient cohort.

## Results

We report results of a multicenter observational study conducted by the BETRnet consortium at 6 participating academic medical centers. Between March 2021 and April 2023, 360 patients were enrolled and 300 (83%, median age of 61±13, Table 1) successfully underwent an unsedated esophageal sampling with EsoCheck balloon. Success in administering the EsoCheck device varied from 77% to 92% among the 7 research coordinators who performed the procedure. Participants in trial reported little to no anxiety, pain, or choking, as reflected by scores of 3 or less on a 10-point Likert scale, by respectively 81%, 95%, and 79% of 296 completed surveys (supplementary table 1). Severe gagging during swallowing was the most common complaint, with 47% reporting low-level gagging (scores 3 or below on 10-point Likert scale), and only 24% scoring gagging levels at or above 7. Overall tolerance was excellent (score ≤3) in 72% of cases, and 94% of individuals who participated in esophageal balloon testing considered it acceptable (scores of 7 or less, Supplementary table 1). A majority of patients (94%) would repeat the procedure again if necessary, and would recommend it to others (supplementary table 2).

**Table 1:** Demographic characteristics of evaluable subjects in EsoCheck balloon study. Continuous variables (age) were compared by T-tests and categorical variable by Fisher’s exact test. Percentages do not always add up to 100 due to rounding.

|  |  | Total enrollment | Excluded | Controls | Cases | P value, case vs control |
| --- | --- | --- | --- | --- | --- | --- |
|  |  | 300 | 57 | 155 | 88 |  |
| Age, median |  | 61±14 | 58±12 | 57±14 | 68±10 | <0.0001 |
| Race | white | 89% | 88% | 88% | 92% | 0.3891 |
|  | African American | 6% | 4% | 8% | 5% |  |
|  | Other/unknown | 5% | 9% | 5% | 3% |  |
| Sex | male | 55% | 56% | 41% | 78% | <0.0001 |
|  | female | 45% | 44% | 59% | 22% |  |
| Smoking | Smoker | 48% | 39% | 42% | 65% | 0.0008 |
|  | Nonsmoker | 52% | 61% | 57% | 35% |  |
|  | Unknown | 0% | 0% | 1% | 0% |  |

Eighty six percent (259 of 300 collected samples) yielded adequate DNA amount to perform the EsoGuard assay. As per pre-specified criteria, six individuals with a history of BE were excluded from analysis as no BE was detected on the study endoscopy. Also, as per pre-specified criteria, 10 individuals with nondysplastic intestinal metaplasia of < 1 cm in length at the gastroesophageal junction (ultrashort BE) or intestinal metaplasia of with intestinal metaplasia of the cardia were segregated for separate analysis [11]. The 243 evaluable individuals included 155 controls free of intestinal metaplasia, and 88 analytic cases, 70 with BE (31 non-dysplastic (ND), 14 low grade dysplasia (LGD), 23 high grade dysplasia (HGD), 2 indefinite for dysplasia (IND)), and 18 with EAC or junctional cancers (Table 2). While having similar racial composition, controls were younger than cases, with median age of 57 vs 68 (p<0.0001), and included more females (Table 1), similar to previous studies, and reflecting the expected epidemiology for the esophageal neoplasia.

**Table 2:** EsoGuard performance in room temperature preservative-stored EsoCheck balloon samples. *VIM* and *CCNA1* gene methylation (EsoGuard assay panel) was determined in DNA samples from non-endoscopic EsoCheck balloon sampling of the distal esophagus from: Unaffected controls (individuals with GERD, erosive esophagitis, or no pathology detected during endoscopy); and from cases of Nondysplastic Barrett’s Esophagus (NDBE); Barret’s Esophagus with Low-Grade or indefinite for Dysplasia (LGD and indefinite); Barrett’s Esophagus with High-Grade Dysplasia (HGD); Esophageal adenocarcinoma and/or junctional cancer of the esophagus (EAC). Samples were scored as *VIM* methylated for mVIM >1.0%, and as *CCNA1* methylated for mCCNA1 >0.5%. Samples were positive for the panel of mCCNA1 plus mVIM if either marker tested positive. Entries indicate percent sensitivity or specificity (%) and total number of individuals tested (N).

|  | EsoGuard panel |  |  |  |
| --- | --- | --- | --- | --- |
|  | VIM | CCNA | VIM+CCNA | No. of Patients |
| Specificity | 86% | 92% | 85% | 155 |
| Overall Sensitivity | 78% | 73% | 85% | 88 |
| Sensitivity NDBE | 74% | 68% | 84% | 31 |
| Sensitivity LGD and indefinite | 75% | 63% | 81% | 16 |
| Sensitivity HGD | 74% | 70% | 78% | 23 |
| Sensitivity EAC | 94% | 94% | 100% | 18 |

(10 cases were excluded due to intestinal metaplasia of the cardia or due to nondysplastic intestinal metaplasia of < 1 cm in length at the gastroesophageal junction; Three of the 10 ultrashort/CIM cases were EsoGuard positive.)

In these room-temperature-preserved balloon samples, the two-marker EsoGuard assay, consisting of mVIM and mCCNA1, showed overall sensitivity for detecting all cases of 85% (95% CI= 0.78-0.93) and specificity for controls of 85% (95% CI=0.79-0.90), a performance highly similar to that found previously in frozen samples (Table 2). While the two marker EsoGuard combination showed overall 85% sensitivity for detecting all cases, this included 100% sensitivity for detecting cancers, 78% for detecting HGD, and 84% sensitivity for detecting NDBE (Table 2). While nondysplastic intestinal metaplasias of < 1 cm were prespecified as excluded from analysis, we did include 3 analytic cases of HGD with lesions of < 1 cm of size. Restricting analysis only to cases with BE family lesions of ≥ 1 cm in size would have increased sensitivity to 86% for all cases and to 80% for detecting HGD.

The EsoCheck balloon samples approximately 5 cm of the distal esophagus. While long-segment BEs (non-dysplastic and dysplastic) were detected with higher sensitivity than the short-segment BEs of only 1 - <3 cm in length (88% vs 76% respectively, table 3), the difference was not statistically significant (p= 0.2300). However, 20 of the 33 short-segment BE were non-circumferential lesions, 18 classified as C0M1 and 2 as C0M2. Overall, the results support that the EsoCheck-EsoGuard technology is sensitive to detecting even small amount of aberrantly-methylated cells in short-segment lesions.

**Table 3:** Effect of BE length on EsoGuard performance in room temperature preservative-stored EsoCheck balloon samples. *C*ases of Nondysplastic Barrett’s Esophagus (NDBE); Barret’s Esophagus with Low-Grade or indefinite for Dysplasia (LGD and indefinite); and Barrett’s Esophagus with High-Grade Dysplasia (HGD) were classified based on the length of the BE segment as Long-segment BE (LSBE) if they had the length greater or equal to 3 cm on endoscopy, or Sort-segment BE (SSBE), if the length of the BE was 1-3 cm. Samples were scored as *VIM* methylated for mVIM >1.0%, and as *CCNA1* methylated for mCCNA1 >0.5%. Samples were positive for the EsoGuard panel of mCCNA1 plus mVIM if either marker tested positive. Entries indicate percent sensitivity or specificity (%) and total number of individuals tested (N).

|  | LSBE |  |  |  | SSBE |  |  |  | P value, LSBE vs SSBE |
| --- | --- | --- | --- | --- | --- | --- | --- | --- | --- |
|  | VIM | CCNA | VIM+ CCNA | No. of Patients | VIM | CCNA | VIM+ CCNA | No. of Patients |  |
| Overall Sensitivity | 88% | 76% | 88% | 33 | 62% | 59% | 76% | 37 | 0.2300 |
| Sensitivity NDBE | 75% | 67% | 75% | 12 | 74% | 68% | 89% | 19 | 0.3499 |
| Sensitivity LGD and indefinite | 100% | 80% | 100% | 10 | 33% | 33% | 50% | 6 | 0.0357 |
| Sensitivity HGD | 91% | 82% | 91% | 11 | 58% | 58% | 67% | 12 | 0.3168 |

(SSBE HGD includes 3 cases with < 1 cm of disease, of which 2 were detected as methylation positive.)

While the females were under-represented in the case group, due to disease being more prevalent in males, sensitivity for disease detection was similar in both males and females (86% vs 84%, table 4). A positive or negative history of smoking also had no significant effect on sensitivity for disease detection (table 5). Specificity rate was similar in male (84%) and female (85%) controls and was also not affected by the patient’s smoking history (Tables 4 and 5).

**Table 4:** Patient gender versus EsoGuard performance in room temperature preservative-stored EsoCheck balloon samples. Unaffected controls (individuals with GERD, erosive esophagitis, or no pathology detected during endoscopy); and cases of Nondysplastic Barrett’s Esophagus (NDBE); Barrett’s Esophagus with Low-Grade or indefinite Dysplasia (LGD and indefinite); and Barrett’s Esophagus with High-Grade Dysplasia (HGD) were classified based on their biological sex. Samples were scored as *VIM* methylated for mVIM >1.0%, and as *CCNA1* methylated for mCCNA1 >0.5%. Samples were positive for the panel of mCCNA1 plus mVIM if either marker tested positive. Entries indicate percent sensitivity or specificity (%) and total number of individuals tested (N).

|  | Male |  |  |  | Female |  |  |  | P value, male vs female |
| --- | --- | --- | --- | --- | --- | --- | --- | --- | --- |
|  | VIM | CCNA | VIM+ CCNA | No. of Patients | VIM | CCNA | VIM+ CCNA | No. of Patients |  |
| Specificity | 86% | 95% | 84% | 63 | 87% | 90% | 85% | 92 | 1.0000 |
| Overall Sensitivity | 80% | 71% | 86% | 69 | 74% | 79% | 84% | 19 | 1.0000 |
| Sensitivity NDBE | 68% | 58% | 79% | 19 | 83% | 83% | 92% | 12 | 0.6236 |
| Sensitivity LGD and indefinite | 75% | 63% | 81% | 16 | - | - | - | 0 | N/A |
| Sensitivity HGD | 79% | 74% | 84% | 19 | 50% | 50% | 50% | 4 | 0.1937 |
| Sensitivity EAC | 100% | 93% | 100% | 15 | 67% | 100% | 100% | 3 | 1.0000 |

**Table 5:** Effect of smoking on EsoGuard performance in room temperature preservative-stored EsoCheck balloon samples. Unaffected controls (individuals with GERD, erosive esophagitis, or no pathology detected during endoscopy); and cases of Nondysplastic Barrett’s Esophagus (NDBE); Barret’s Esophagus with Low-Grade or indefinite Dysplasia (LGD and indefinite); and Barrett’s Esophagus with High-Grade Dysplasia (HGD) were classified based on their self-reported smoking history as smokers (current or former), and non-smokers (have never smoked). One individual with unknown/unreported smoking history was excluded from this analysis. Samples were scored as *VIM* methylated for mVIM >1.0%, and as *CCNA1* methylated for mCCNA1 >0.5%. Samples were positive for the panel of mCCNA1 plus mVIM if either marker tested positive. Entries indicate percent sensitivity or specificity (%) and total number of individuals tested (N).

|  | Smokers |  |  |  | Non-smokers |  |  |  | P value, smokers vs non-smokers |
| --- | --- | --- | --- | --- | --- | --- | --- | --- | --- |
|  | VIM | CCNA | VIM+CCNA | No. of Patients | VIM | CCNA | VIM+CCNA | No. of Patients |  |
| Specificity | 89% | 91% | 86% | 65 | 84% | 93% | 83% | 89 | 0.6591 |
| Overall Sensitivity | 82% | 77% | 86% | 57 | 71% | 65% | 84% | 31 | 0.7642 |
| Sensitivity NDBE | 75% | 69% | 81% | 16 | 73% | 67% | 87% | 15 | 1.0000 |
| Sensitivity LGD and indefinite | 91% | 73% | 91% | 11 | 40% | 40% | 60% | 5 | 0.2143 |
| Sensitivity HGD | 75% | 69% | 75% | 16 | 71% | 71% | 86% | 7 | 1.0000 |
| Sensitivity EAC | 93% | 100% | 100% | 14 | 100% | 75% | 100% | 4 | 1.0000 |

When the two methylated DNA markers are considered individually, sensitivity for detecting all cases was 78% for mVIM alone, and 73% for mCCNA1 alone (Table 2). While the two marker EsoGuard combination showed overall 85% specificity in normal controls, mCCNA1 showed a highest 92% specificity, while mVIM, exhibited 86% specificity.

## Discussion

Our previous studies have demonstrated that the encapsulated balloon device successfully sampled the distal esophagus with excellent tolerability and acceptability and, the two-marker panel of mVIM plus mCCNA1 demonstrated high sensitivity and specificity for detecting BE in NGS-based bisulfite sequencing assay [11]. The initial study contained only 50 cases and 36 control patients, was performed in a research lab, and relied on snap-freezing the EC balloon samples immediately after collection, in order to preserve the DNA and the methylation signature of the sample. The wider clinical implementation of this test depended, in part, on the ability to preserve the DNA methylation at ambient temperature, facilitating the shipment of the samples to a central lab for testing.

The current study shows that the new iteration of EsoGuard/Esocheck has been successfully adapted to a room temperature preservative, and has been successfully performed in a CLIA certified laboratory, enabling point of care office-based testing. The EC procedure continues to be well-tolerated by the patients, and the new-generation EG test detects non-dysplastic BE, dysplastic BE, and cancer with high sensitivity and specificity, replicating the findings of the initial pilot study of this technology in a larger group of patients. We additionally note the recent report of a 1483 patient cohort, that, with further training and further technical enhancements to sample processing, achieved a 98% success rate in EC administration, with 97% of samples yielding adequate DNA for EG assay [12].

EG/EC is currently the only commercially available non-endoscopic screening test for BE in the US, and this study shows that the test can be practical for clinical implementation, and viable for office-based screening, enabling the more wide-spread screening for BE as recommended in the most recent ACG Guideline and AGA Clinical Update. Larger studies validating population-based implementation, physician adoption, and patient acceptance of BE and EC screening with EG/EC are needed.

## Materials and Methods

### Study Design

The study protocol was approved by Institutional Review Boards for Human Subject Investigation at all 6 participating medical centers: University Hospitals Case Medical Center, Cleveland Clinic, Mayo Clinic, The Johns Hopkins Medical Institutions, Washington University School of Medicine, University of North Carolina-Chapel Hill. Clinical trial registration number at ClinicalTrials.gov is NCT00288119 for the non-endoscopic balloon trial. Subjects referred for outpatient EGD were approached for study participation. Consent for obtaining esophageal brushings and biopsies for research was obtained from subjects prior to their EGD. Cases were classified as subjects with newly diagnosed BE, those undergoing surveillance of BE, or those with a new diagnosis of esophageal or gastroesophageal junctional adenocarcinoma undergoing an endoscopic procedure. BE was defined according to current ACG guidelines as at least one cm of endoscopically visible columnar mucosa in the distal esophagus with intestinal metaplasia confirmed on histology [9]. Lesions < 1 cm or biopsies of an irregular Z-line with non-dysplastic IM were classified as intestinal metaplasia of the cardia and excluded. All dysplastic lesions including LGD or HGD regardless of length were included. Control subjects had no endoscopic evidence of BE and no histological evidence of intestinal metaplasia if a clinical biopsy was obtained from either the distal esophagus or gastroesophageal junction.

The overall study is a non-randomized observational study. Study size was not pre-specified, and results are reported for esophageal balloon samples accrued from March 2021 to April of 2023. Subjects were excluded from the primary analysis and reported separately if they had one of the following: no endoscopy performed at the time of balloon swallow, history of prior esophageal ablation therapy, history of intestinal metaplasia (IM), but no IM found at the time of balloon swallow, evidence of Gastric intestinal metaplasia (GIM), intestinal metaplasia of the cardia (CIM), or stomach IM. The primary endpoint of detection of BE and related progressed lesions was pre-specified prior to study initiation. All laboratory samples were assayed by technicians in a CLIA-certified lab blinded to the clinical status of the subjects. Patient demographics and clinical diagnosis were collected by a separate group of study site coordinators who were blinded to the methylation status of the samples.

### Non-endoscopic esophageal sampling via a balloon device

EsoCheck device was used as previously described [11] by nurses trained in balloon sampling procedure. Subjects referred for outpatient EGD at participating BETRnet study location were approached for study participation. Patients underwent un-sedated distal esophageal sampling with the balloon device prior to scheduled EGD. Following the procedure, each patient filled out a standardized tolerance and acceptance questionnaire [13], Supplementary document 1. After the procedure, balloons were re-inflated, cut off from the capsule, and immediately dropped into vials containing a preservative storage buffer (Lucid Diagnostics). The vials were tightly capped and shaken to ensure full submersion of the capsule and sample preservation. Samples were shipped to the Lucid Diagnostics CLIA-compliant lab for processing. DNA was extracted and stored at -70C^0^ until analyzed by the EsoGuard assay implementation of bisulfite sequencing for detection of aberrant methylation in the vimentin and CCNA1 genomic loci (mVim and mCCNA1, respectively), as previously described [11]. When, for quality control purposes, a sample was run more than once, the results of the replicate runs were averaged.

### Statistical methods

Between-group comparisons of continuous variables were performed using unpaired t test (for two groups). Fisher’s exact test was used for comparison of demographic composition of cases/controls. Confidence interval for proportions was calculated using the Willson score interval.

## Data Availability

All data produced in the present study are available upon reasonable request to the authors

## ACKNOWLEDGEMENTS

Supported by NIH grants P01CA269019, P50CA150964, U54CA163060, UO1CA152756, U01CA271867, UL1TR000439, and by Lucid Diagnostics.

We would like to thank Wendy Brock, BSN, RN (University Hospitals Cleveland Medical Center), Beth Bednarchik, BSN, RN, CCRP (University Hospitals Cleveland Medical Center), Vidhi Patel, MD (Cleveland Clinic), Rajesh Guptha Bhaskaran, RN, MS (Case Western Reserve University), Thomas Hollander, RN, MS, BSN, CCRP (Washington University at St Louis School of Medicine), Hilary Cosby, RN, CGRN (Johns Hopkins Medical Institute), Megan Dianne Ramsey, BS (University of North Carolina), Lama Moussa, BS (University of North Carolina), and Ramona Lansing, RN (Mayo Clinic Rochester) for their efforts in recruiting patients, administering the EC/EG test, collecting data, and coordinating the research.

