## Supplemental Tables for "NON-ENDOSCOPIC ESOPHAGEAL SAMPLING DEVICE AND BIOMARKER PANEL FOR DETECTION OF BARRETT’S ESOPHAGUS (BE) AND ESOPHAGEAL ADENOCARCINOMA (EAC)"

**Supplementary document 1. Balloon trial questionnaire**

**PROCEDURE TOLERABILITY AND ACCEPTABILITY**

**(Complete after the capsule balloon test)**

Next, we want to ask you some questions about **YOUR EXPERIENCE** with the Capsule Balloon procedure. Please mark only one number for each question by filling in the circle completely.

1. Your level of anxiety, nervousness or worried feelings that you experienced about having the capsule test.
   **No worries** **I was terrified**
   O 1 O 2 O 3 O 4 O 5 O 6 O 7 O 8 O 9 O 10

2. The level of pain that you experienced with the procedure.
   **No pain** **Most severe pain**
   O 1 O 2 O 3 O 4 O 5 O 6 O 7 O 8 O 9 O 10

3. The level of gagging or retching that you experienced with the procedure.
   **No gagging** **Most severe gagging**
   O 1 O 2 O 3 O 4 O 5 O 6 O 7 O 8 O 9 O 10

4. The level of choking you experienced during the procedure.
   **No choking** **Most severe choking**
   O 1 O 2 O 3 O 4 O 5 O 6 O 7 O 8 O 9 O 10

5. The level of overall tolerance of the procedure.
   **No difficulty** **Severely intolerable**
   O 1 O 2 O 3 O 4 O 5 O 6 O 7 O 8 O 9 O 10

6. We would appreciate **written comments** regarding what was the most difficult part of the capsule balloon procedure:

   ________________________________________

   ________________________________________

**Acceptability Of Screening Test**

**Please indicate the extent to which you agree or disagree with each of the following statements. Use a scale of 1 to 5, with 5 being Strongly Agree and 1 being Strongly Disagree. If an item is not related to your care choose N/A.**

| | Strongly Disagree (1) | Somewhat Disagree (2) | Neutral (3) | Somewhat Agree (4) | Strongly Agree (5) | N/A |
|---|---|---|---|---|---|---|
| I would undergo the balloon test again if needed for further care. | O | O | O | O | O | O |
| I would recommend this screening balloon test to family and friends. | O | O | O | O | O | O |

**Supplementary Table 1: EC balloon procedure tolerability summary.** Tolerability was scored on 10-point Likert scale, with 1 representing the best possible outcome, and 10 representing the worst outcome. 296 patients have completed the questionnaire. For the analysis purpose, the 10-point scale was reduced to three categories: acceptable, intermediate, and unacceptable. The percentage of patients in each group is reported in the table.

| | **Level of anxiety, nervousness, or worried feelings that you experienced about having the capsule test** | **Level of choking you experienced during the procedure** | **Level of gagging or retching that you experienced with the procedure** | **Level of overall tolerance of the procedure** | **Level of pain that you experienced with the procedure** |
|---|---|---|---|---|---|
| Levels of 3 or below (acceptable) | 81% | 79% | 47% | 72% | 95% |
| Levels 4-6 (intermediate) | 15% | 15% | 29% | 21% | 5% |
| Levels of 7 of more (unacceptable) | 4% | 6% | 24% | 6% | 0% |

**Supplementary Table 2: EC balloon procedure acceptability summary.** Acceptability was scored on 5-point Likert scale, with 1 representing the strongest disagreement, and 5 representing the strongest agreement. 296 patients have completed the questionnaire. For the analysis purpose, the scores of 3 and above were considered acceptable, and scores of 2 or below were considered unacceptable. The percentage of patients in each group is reported in the table.

| | **Would prefer the balloon test to upper endoscopy if I need Just ONE test for screening again** | **Would recommend this screening balloon test to family and friends** | **Would undergo the balloon test again if needed for further care** |
|---|---|---|---|
| Scores of 3- 5 (acceptable) | 87% | 94% | 94% |
| Scores of 1 or 2 (unacceptable) | 13% | 6% | 6% |
